# Detecting Patient Position Using Bed-Reaction Forces and Monitoring Skin-Bed Interface Forces for Pressure Injury Prevention and Management

**DOI:** 10.1101/2022.03.15.22272323

**Authors:** Nikola Pupic, Sharon Gabison, Gary Evans, Geoff Fernie, Elham Dolatabadi, Tilak Dutta

## Abstract

Pressure injuries are largely preventable, yet they affect one in four Canadians across all healthcare settings. A key best practice to prevent and treat pressure injuries is to minimize prolonged tissue deformation by ensuring at-risk individuals are repositioned regularly (typically every 2 hours). However, adherence to repositioning is poor in clinical settings and expected to be even worse in homecare settings.

Our team has designed a position detection system for home use that uses machine learning approaches to predict a patient’s position in bed using data from load cells under the bed legs. The system predicts the patient’s position as one of three position categories: left-side lying, right-side lying, or supine. The objectives of this project were to: i) determine if measuring ground truth patient position with an inertial measurement unit can improve our system accuracy (predicting left-side lying, right-side lying, or supine) ii) to determine the range of transverse pelvis angles (TPA) that fully offloaded each of the great trochanters and sacrum and iii) evaluate the potential benefit of being able to predict the individual’s position with higher precision (classifying position into more than three categories) by taking into account a potential drop in prediction accuracy as well as the range of TPA for which the greater trochanters and sacrum were fully offloaded.

Data from 18 participants was combined with previous data sets to train and evaluate classifiers to predict the participants’ TPA using four different position bin sizes (∼70°, 45°, ∼30°, and 15°) and the effects of increasing precision on performance, where patients are left side-lying at -90°, right side-lying at 90° and supine at 0°). A leave-one-participant-out cross validation approach was used to select the best performing classifier, which was found to have an accuracy of 84.03% with an F1 score of 0.8399. Skin-bed interface forces were measured using force sensitive resistors placed on the greater trochanters and sacrum. Complete offloading for the sacrum was only achieved for the positions with TPA angles <-90° or >90°, indicating there was no benefit to predicting with greater precision than with three categories: left, right, and supine.

## 1 Introduction

Pressure injuries (PIs), also known as bed sores or pressure ulcers, are largely preventable, yet they affect one in four Canadians across all healthcare settings (1). PIs are thought to be the result of prolonged deformation resulting from tissues being compressed between a support surface and a bony prominence (2); however, on a microscopic level, tissue deformation can occur within minutes (3,4). Deep tissue injuries are more related to the pressure force and superficial skin injuries are more related to the shear force (4).

The deformation disrupts homeostasis at the cellular level, resulting in a positive feedback cycle of inflammation, ischemia, and cell death (3–5). The current practice for treating pressure injuries is to minimize the risk of prolonged deformation by repositioning patients every two hours to allow the compressed tissues to return to their normal state (3–6). Unfortunately, evidence suggests that adherence rates to repositioning schedules are poor in clinical environments (7–9) and are likely to be worse in homecare settings (10).

The cost of PIs to the healthcare system is also enormous, with the estimated cost currently exceeding $26.8 billion in the United States (11). These costs may soon increase as the COVID-19 pandemic has likely exposed more individuals to PI risks while being treated as in-patients in hospitals (3).

To address the need for improved repositioning, Wong et al. (12) have developed a system that uses machine learning to detect the position of a simulated patient in bed based on data from load cells under each bed leg. The proof-of-concept work was able to detect healthy participant position (supine, left-side, or right-side) with 94.2% accuracy (n=20). When Wong et al.’s (12) model was tested on the data collected from nine older adults sleeping in their own beds at home, the accuracy dropped to ∼88.5%. The drop in accuracy was suspected to be due to the large variations of sleeping positions that can be adopted. Additionally, this highlighted the importance of defining which areas of the pelvis are offloaded in different positions.

The primary aim of this work was to improve the performance of our machine learning model using a pelvis-mounted Inertial Measurement Unit (IMU) to provide more accurate ground-truth labels than we previously had using time-lapse images of the individual in bed. The secondary aim was to determine the range of transverse pelvic angles that completely offloads the greater trochanter and the sacrum, where complete offloading indicates there is no force from the bed being applied to the area in question. These results were then used to evaluate the benefit of predicting an individual’s position in bed with finer precision than supine, left, or right. This was achieved by specifying more narrow ranges of angles within which the participant position belonged to (i.e., a position held at 30° would fall into the *20° to 40°* bin instead of the *Right* bin).

The participant’s position was defined by the angle of the pelvis with respect to the bed in the transverse plane, referred to as the transverse pelvic angle (TPA). Left-side, supine, and right-side lying were represented by -90°, 0°, and 90°, respectively. Figure 1 shows a visual representation of the TPA.

**Figure 1.**
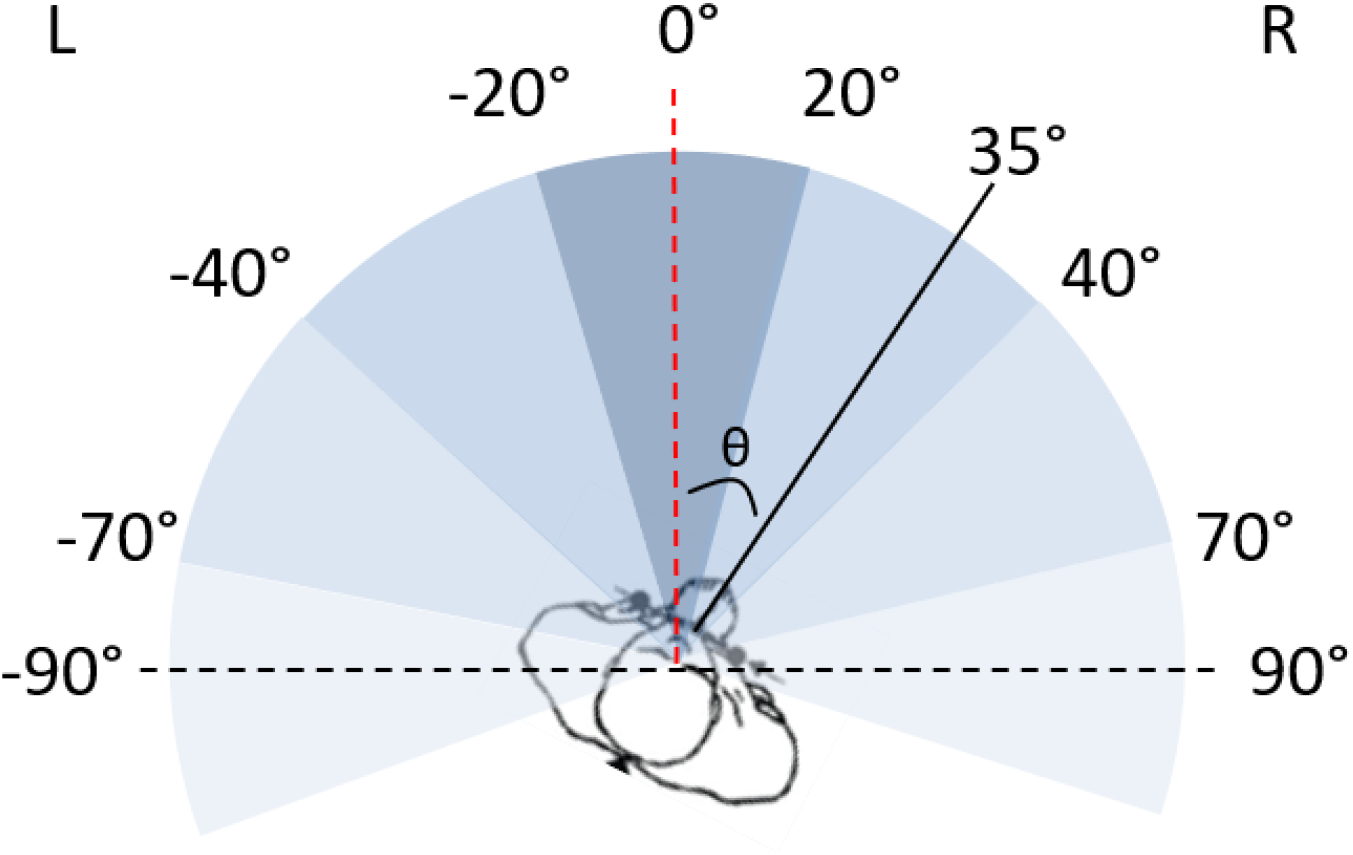
The solid-coloured black line represents an individual’s position on the bed in the transverse plane. This line is perpendicular to the line connecting the left and right anterior superior iliac spine bony landmarks on the individual. The dotted red line represents our 0° (true supine) reference point, and it is defined as the line perpendicular to the surface of the bed. Therefore, the TPA, represented by θ, is defined as the angle between the 0° reference line and the line perpendicular to the anterior-posterior axis of the pelvis.

Figure 2 shows the different bin sizes that were tested to see the effects of increased precision on performance. A total of four different bin sizes were used. For the 15° (Figure 2a) and 45° (Figure 2c) bins, all bins are the same size. However, for the ∼30° (Figure 2b) and ∼70° (Figure 2d) bins, the bin size containing the 0° supine position was adjusted to avoid the bin boundaries coinciding with the positions that participants were asked to adopt.

**Figure 2a.**
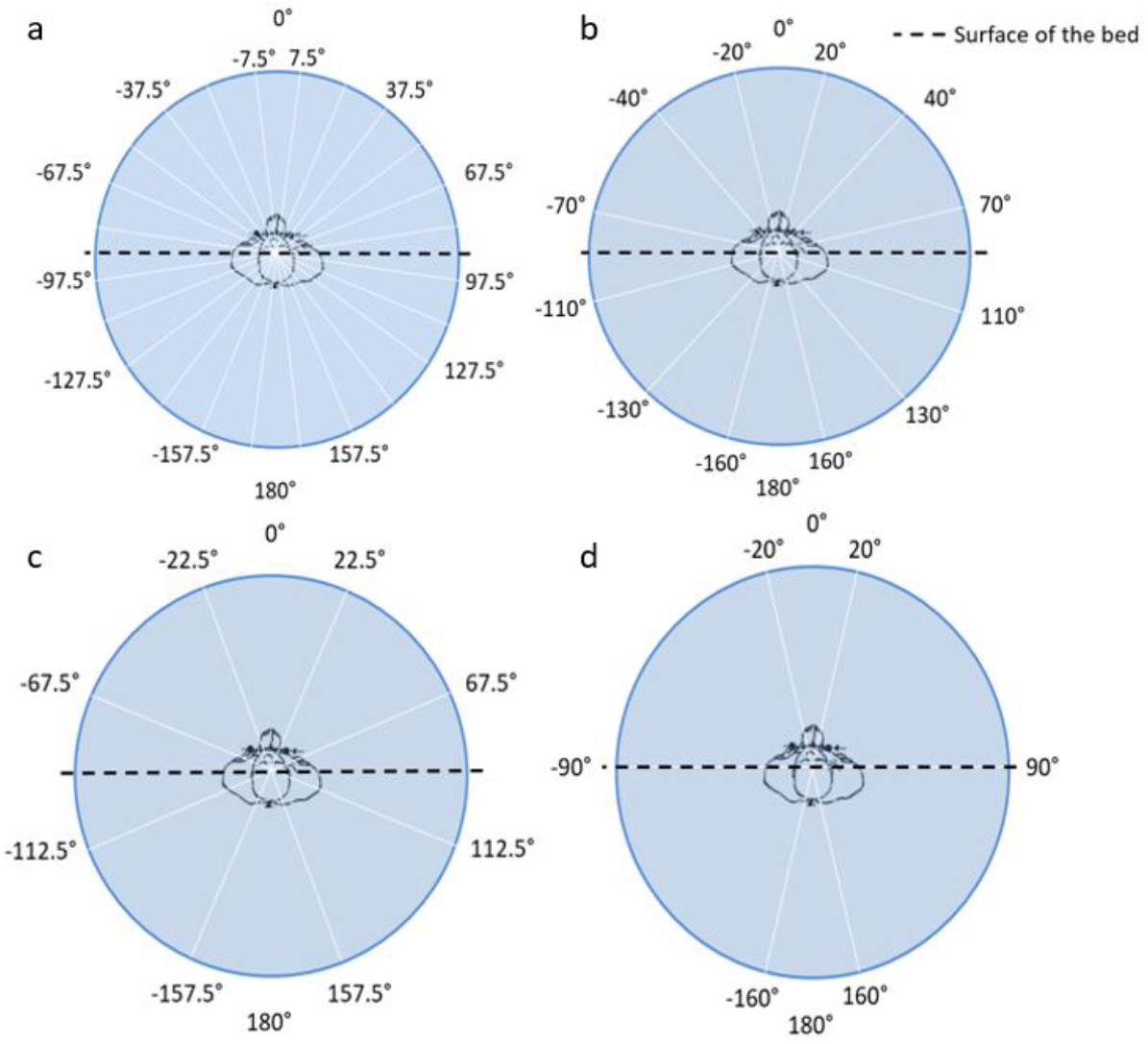
15° bins; **2b**. ∼30° bins; **2c**. 45° bins; **2d**. ∼70° bins

## 2 Methods

### 2.1 Participants

A convenience sample of 20 healthy participants (10 males, 10 females) was recruited for this study. Able-bodied participants were included with no existing pressure injuries. All participants provided their informed consent, and the study protocol was reviewed by the Research Ethics Board of University Health Network.

### 2.2 System Setup

The instrumentation was set up in a similar manner to Wong et al. (12). Data was collected in CareLab, a simulated patient care environment located within Toronto Rehabilitation Institute (TRI), using a Carroll hospital bed (Carroll Hospital Group, Kalamazoo, MI). Single-axis load cells comprised of four load sensors (model DLC902-30KG-HB, Hunan Detail Sensing Technology, Changsha, Hunan, China) arranged in a full Wheatstone bridge circuit were placed under each of the four wheels of the bed. The load cell signals were amplified, filtered, and converted from analog to digital using a signal conditioner (GEN 5, AMTI, Watertwon, MA) configured for 5.0 VDC excitation and a gain of 500 for each channel. NetForce software (version 3.5.2, AMTI, Watertown, MA) running on a laptop PC (Thinkpad T520, Lenovo, Hong Kong, China, 2.5 GHz Intel Core i5 CPU, 4 GB of RAM) was used to collect the load cell data at 50 Hz with 16-bit resolution. A camera was also positioned above the bed to capture ground truth video data of the participant positions.

Participants were fitted with three IMUs (Shimmer3, Shimmer Sensing, Dublin, Ireland) – one around the chest, one around the pelvis, and one around the arm in order to collect ground truth data for the sternal angle, pelvic angle, and heart rate, respectively.

The IMUs were connected to a laptop (Aspire 5, Acer, Xizhi, New Taipei, Taiwan, 1.8GHz Intel Core i7 CPU, 12GB of RAM) via Bluetooth to ConsensysPRO (version 1.6.0, Consensys, Dublin, Ireland) to collect data, visualize real-time transverse trunk and pelvic angles, and choose the sampling frequency (256 Hz).

Participants were also fitted with three FSRs (model RP-S40-ST, Hilitand) in order to collect offloading data. The FSRs were connected to an Arduino board, which was connected to a laptop (Aspire 5, Acer, Xizhi, New Taipei, Taiwan, 1.8GHz Intel Core i7 CPU, 12GB of RAM) using the Arduino software (Arduino, version 1.8.13, Boston, MA). Three LED lights (one for each sensor) were set up to indicate that the FSRs were recording properly.

### 2.3 Data Collection

The data was collected in two phases: a) the primary phase, where participants were instructed to cycle through a series of 11 unique positions at 0°, ±15°, ±30°, ±45°, ±60°, and ± 90°; and b) the random phase, where participants could assume any position they wanted to from -180° to +180° to account for the wide range of positions that can be adopted in bed. In addition, participants were asked to assume one prone position.

All primary and random phase positions were held for three minutes. Figure 3 shows order of positions, including the intermediate positions (defined as positions held in between each three-minute hold,) that were held for one minute. The intermediate positions served as a way of standardizing the way pillows were inserted and removed for the primary phase.

**Figure 3.**
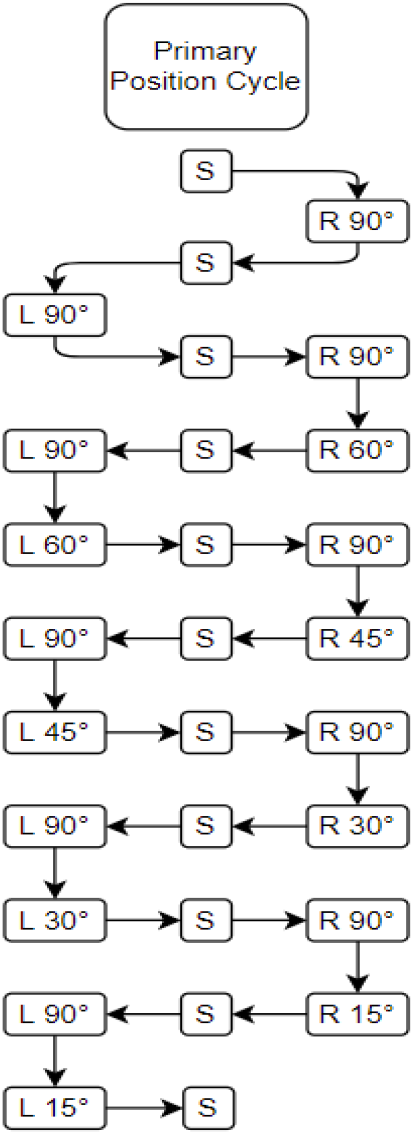
The 11 different positions adopted by patients during primary testing and the order of positions, including Intermediate holds at supine, R90° and L90°.

Data from two participants was removed from the data set due to equipment malfunction.

### 2.4 Data Supplementation

Additional data was incorporated into the training set to increase the size of the data set. This additional data was only used to train machine and deep learning classifiers for the detection of supine, left, or right positions. The total data set used included 20,520 observations of which 2,963 observations were from data collected in this study; 4,909 observations were from the data collected by Wong et al. (12); and 12,918 observations were from data collected in the home environment by Gabison et al. (13).

### 2.5 Data Processing

#### Load Cell Data

Load cell signals were exported from Netforce and processed offline using MATLAB 2020a. The data was manually segmented into trials by removing sections where the participants were changing positions. Next, the center of mass of the bed-patient system was calculated using equations 1 and 2 below where CoM_x and CoM_y refer to the center of mass in the x (parallel to the short axis or width of the bed) and y (parallel to the long axis or length of the bed) directions, respectively. The data processing will be performed in the same manner as the study by Wong et al (12). Below is an explanation of how the data processing was executed, where LH

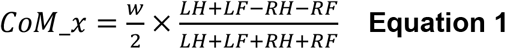

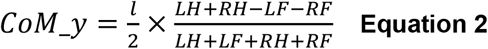

and RH correspond to the vertical forces measured by left and right sensors at the head of the bed respectively, LF and RF corresponds to the vertical forces measures by the left and right sensors at the foot of the bed respectively, and *l* and *w* refer to the distances between the load cells. To isolate the changes in the CoM signals associated with respiration, CoM_x and CoM_y signals were low pass filtered with personalized Chebyshev Type II filters. This filter was applied using MATLAB’s filtfilt function (ensuring zero-phase shift) to obtain CoM_resp_x and CoM_resp_y. The times when maxima (tmax) and minima (tmin) occurred in the CoM_resp_x and CoM_resp_y signals were found by finding zero crossings for the first derivative of each signal. These times correspond with the end of each exhalation and inhalation respectively (14). The angle of the principal axis of the ellipsoid traced by the resultant CoM_resp signal relative to the positive x axis (positive angle measured clockwise) was calculated using equation 3 for each tmax and subsequent tmin.

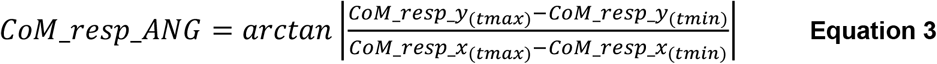

Finally, components of the signal that captured changes resulting from the cardiac cycle (rmsPulse) were isolated. MATLAB’s filtfilt function was used to bandpass filter the sum of the LH and RH signals using a personalized equiripple finite impulse response filter. Each data point used for training/testing the machine learning and deep learning classifiers was the average of a 45 s moving window (2250 observations) with a new value computed by shifting the window by 15 s. Since each pose was maintained for approximately 3 min, roughly 10 data points were calculated for each pose with each participant. Missing data from one participant was interpolated based on values from the same window.

#### IMU Data

The ground truth data was classified into one of three positions: right-side lying, left-side lying, or supine. The classifications were made based on a combination of Euler angles generated from the IMU data (data collected during this study), video data (data collected by Wong et al. (12)), and time-lapsed images (data collected in a home environment and classifications performed by 3 independent and blinded raters). The IMU data was further classified four more times using the generated Euler angles, once for each of the different bin sizes, to allow for more precise TPA detection.

#### FSR Data

Only FSR data from the primary phase was used for this analysis as the positions were consistent between all participants. The FSR data was manually annotated by the author to assign position codes using the video data as ground truth guidance. A total of 865,234 observations were collected. Sensor malfunctions (incorrect readings where the sensor reported maximum values when there was no force placed on it or no force when there was forced placed on it) and transitions (readings that occurred while positions were being changed and that have no use) resulted in 84,905 and 36,487 observations being removed, respectively. The final reported data set contained 743,932 observations.

The FSR data was converted from ADC values to Resistance (Kiloohms) using Equation 4 and then from Resistance (Kiloohms) to Force (grams) using Equation 5.

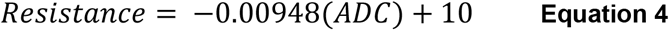

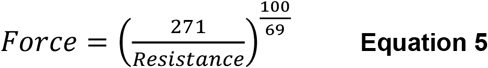

As the FSRs were attached to participants using tape, there was a constant force (approximately 120g) present on the sensors. This was referred to as the “tape bias” and was defined as the lowest recorded force value present. The tape bias was subtracted from the force measurements and the values were normalized by dividing all the recordings for a participant by the maximum achieved force reading for that participant. As such, all force values were reported as the percentage of maximum force at each position for the participant.

### 2.6 Feature Selection

The same features that Wong et al. (12) used to achieve an accuracy of 94.2% with their in-lab participant study were used. These features were extracted from the load cell data (Table 1).

**Table 1.**
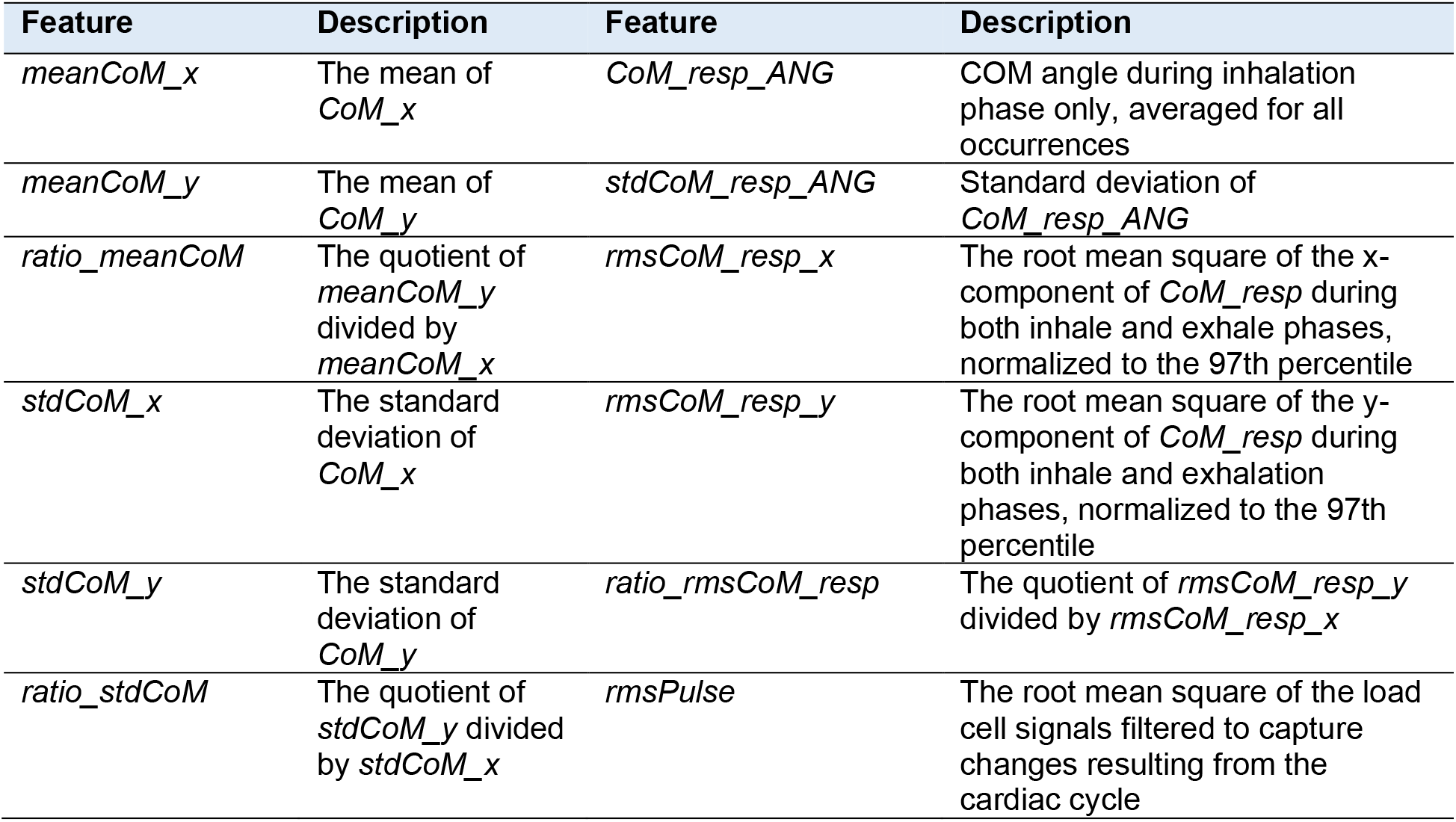
Features extracted by Wong et al.

| Feature | Description | Feature | Description |
| --- | --- | --- | --- |
| *meanCoM_x* | The mean of *CoM_x* | *CoM_resp_ANG* | COM angle during inhalation phase only, averaged for all occurrences |
| *meanCoM_y* | The mean of *CoM_y* | *stdCoM_resp_ANG* | Standard deviation of *CoM_resp_ANG* |
| *ratio_meanCoM* | The quotient of *meanCoM_y* divided by *meanCoM_x* | *rmsCoM_resp_x* | The root mean square of the x-component of *CoM_resp* during both inhale and exhale phases, normalized to the 97th percentile |
| *stdCoM_x* | The standard deviation of *CoM_x* | *rmsCoM_resp_y* | The root mean square of the y-component of *CoM_resp* during both inhale and exhalation phases, normalized to the 97th percentile |
| *stdCoM_y* | The standard deviation of *CoM_y* | *ratio_rmsCoM_resp* | The quotient of *rmsCoM_resp_y* divided by *rmsCoM_resp_x* |
| *ratio_stdCoM* | The quotient of *stdCoM_y* divided by *stdCoM_x* | *rmsPulse* | The root mean square of the load cell signals filtered to capture changes resulting from the cardiac cycle |

### 2.7 Machine Learning Approach

A two-part hierarchical classification approach was used (Figure 4), similar to the one used by Liang et al. (15). Level 1 was trained on the final data set and tested only on the new data to predict either supine, left, or right. After this classification, the new data was divided into three separate smaller data sets based on its IMU ground truth label (either supine, left, or right). The data from each of the three positions was fed into its respective model for Level 2 classification, where the model specified a more precise bin of angles that each position belonged to. Level 2 classification was repeated four times, once for each bin size (15°, ∼30°, 45°, ∼70°).

**Figure 4.**
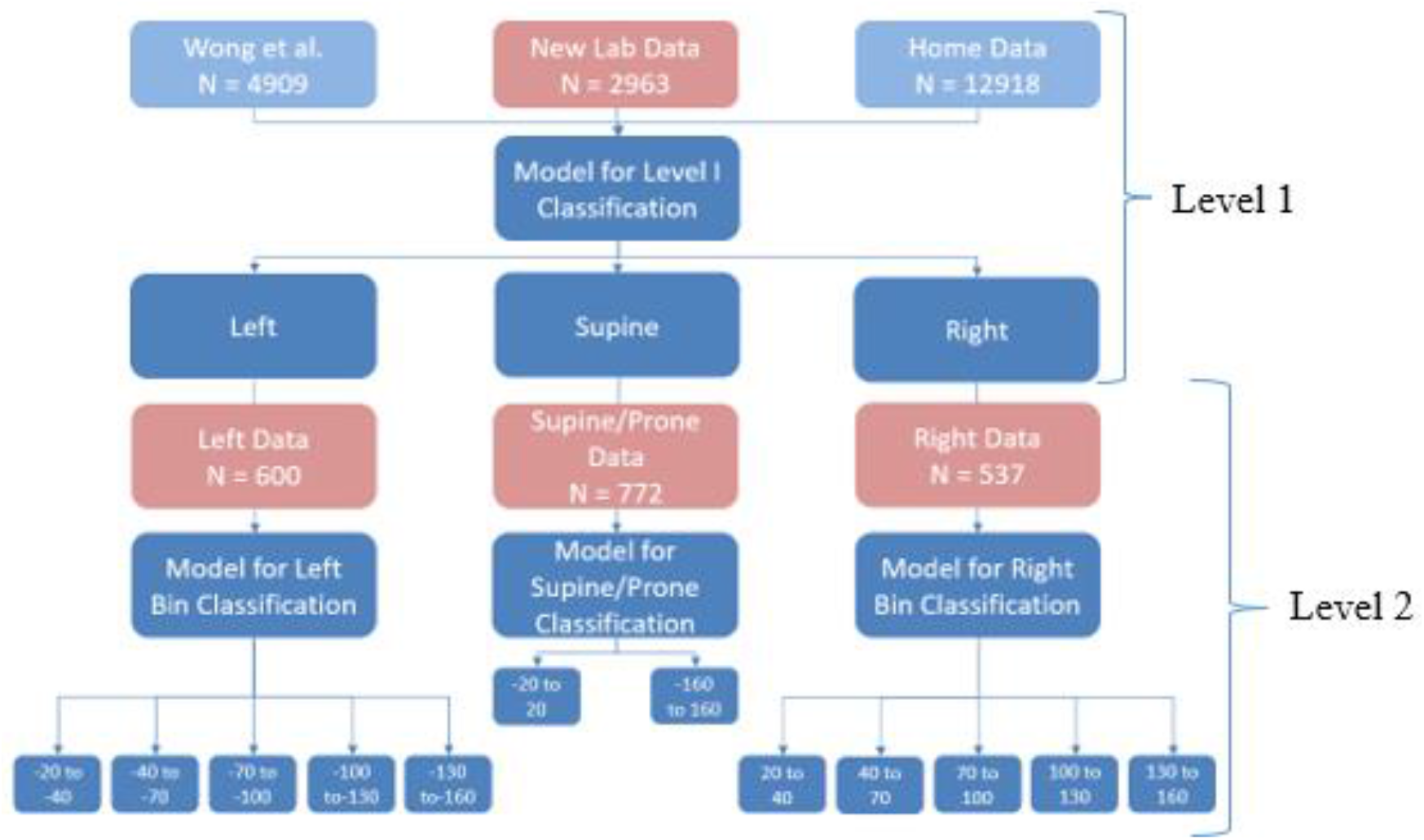
Flow diagram of how the two levels of hierarchical classification work together, where the second level shows the example of a bin size of 30°.

Table 2 describes how the final data set was used for the classification tasks.

**Table 2.** Table describing the different data sources and how they were used.

| Data Source | Data Specifications | Data Set Size (samples) | IMU | Data Use | Hierarchy Level |
| --- | --- | --- | --- | --- | --- |
| Wong et al. (Lab Data) | 20 healthy participants | 4909 | No | Training | 1 |
| Home Data collected by our team | 9 healthy participants collected at home and 1 healthy participant collected in a sleep lab | 12918 | No | Training | 1 |
| New lab data collected for this study | 18 healthy participants | 2963 | Yes | Training & Testing | 1 and 2 |

#### Leave-One-Participant-Out

A leave-one-participant-out cross validation approach was used to evaluate the accuracy of the classifier, while maximizing the number of training observations. Using this method, a classifier was trained on a data set that incorporated 17 participants and tested on the one excluded participant. This procedure was repeated 18 times, once for each participant. The overall performance measures were estimated from the averaged errors for each individual test sample.

#### Incremental Learning

Incremental learning was used to evaluate the potential of the classifier to adapt to the left-out participant. The classifier was trained using different percentages of the left-out participant’s data (c = 0, 10, 20, 30%). To maintain a uniform test set, the left-out participant’s data was split into a 30% incremental learning set, from which different c values were added to the training set, and a 70% test set.

#### Machine Learning Classifiers

Table 3 shows a list of models used in both Level 1 and 2 classifications. For Level 1, both machine and deep learning models were used. For Level 2, only machine learning models were used as there was not enough data to warrant the use of deep learning.

**Table 3.** Table describing the models created, which level they were used for, and the features they used.

| Model | Level 1 Prediction | Level 2 Prediction | Features Used |
| --- | --- | --- | --- |
| Logistic Regression |  | X | 12 features from Wong et al. |
| Support Vector Machine |  | X | 12 features from Wong et al. |
| Gradient Boosting Classifier | X | X | Some of the 12 features from Wong et al. |
| AdaBoost Classifier | X | X | 12 features from Wong et al. |
| XGBoost Classifier | X | X | 12 features from Wong et al. |
| Light Gradient Boosting Machine Classifier | X |  | 12 features from Wong et al. |
| Multilayer Perceptron (x3) | X |  | 12 features from Wong et al. |
| Recurrent Neural Network: Long Short-Term Memory | X |  | 12 features from Wong et al. & Automatic feature selection |
| Convolutional Neural Network: 1-Dimensional | X |  | 12 features from Wong et al. & Automatic feature selection |

Three different MLP models were constructed to test Level 1 classification, where: MLP 1 was the original model used by Wong et al. []; MLP 2 was a large, hyperparameter tuned model using keras.tuner; and MLP 3 was a simple MLP model. Tables 4, 5, and 6 describe the different model architectures.

**Table 4.** Table describing the architecture of MLP 1, the original MLP model used by Wong et al. in their study. The dropout was set to 0.1 for all instances, the batch size was not specified, and the learning rate was 0.001.

| Layers | Number of Nodes | Activation Function |
| --- | --- | --- |
| Input | 12 | ReLu |
| Fully Connected 1 | 64 | ReLu |
| Dropout 1 | N/A | N/A |
| Batch Normalization 1 | N/A | N/A |
| Fully Connected 2 | 100 | ReLu |
| Dropout 2 | N/A | N/A |
| Batch Normalization 2 | N/A | N/A |
| Output | 3 | Softmax |

**Table 5.** Table describing the architecture of MLP 2, the hyperparameter tuned model. The dropout was set to 0.1 for all instances, the batch size was 128, and the learning rate was 0.000417.

| Layers | Number of Nodes | Activation Function |
| --- | --- | --- |
| Input | 12 | ReLu |
| Fully Connected 1 | 128 | ReLu |
| Dropout 1 | N/A | N/A |
| Batch Normalization 1 | N/A | N/A |
| Fully Connected 2 | 256 | ReLu |
| Dropout 2 | N/A | N/A |
| Batch Normalization 2 | N/A | N/A |
| Fully Connected 3 | 160 | ReLu |
| Dropout 3 | N/A | N/A |
| Batch Normalization 3 | N/A | N/A |
| Fully Connected 4 | 32 | ReLu |
| Dropout 4 | N/A | N/A |
| Batch Normalization 4 | N/A | N/A |
| Output | 3 | Softmax |

**Table 6.** Table describing the architecture of MLP 3, the simplified tuned model. The dropout was set to 0.1 for all instances, the batch size was 32, and the learning rate was 0.001.

| Layers | Number of Nodes | Activation Function |
| --- | --- | --- |
| Input | 12 | ReLu |
| Fully Connected 1 | 12 | ReLu |
| Dropout 1 | N/A | N/A |
| Batch Normalization 1 | N/A | N/A |
| Fully Connected 2 | 6 | ReLu |
| Dropout 2 | N/A | N/A |
| Batch Normalization 2 | N/A | N/A |
| Fully Connected 3 | 4 | ReLu |
| Dropout 3 | N/A | N/A |
| Batch Normalization 3 | N/A | N/A |
| Output | 3 | Softmax |

Tables 7 and 8 describe the architecture of the CNN and RNN, respectively.

**Table 7.** Table describing the architecture of the CNN model. The dropout was set to 0.2, 0.0, 0.05, and 0.45 for the four layers, respectively; the batch size was 16, and the learning rate was 0.00188.

| Layer | Number of Filters | Size/Stride | Padding | Activation Function |
| --- | --- | --- | --- | --- |
| Input | 13 | N/A | N/A | ReLu |
| Convolutional 1 | 32 | 1/2 | Same | ReLu |
| Dropout 1 | N/A | N/A | N/A | N/A |
| Batch Normalization 1 | N/A | N/A | N/A | N/A |
| Convolutional 2 | 32 | 7/4 | Same | ReLu |
| Dropout 2 | N/A | N/A | N/A | N/A |
| Batch Normalization 2 | N/A | N/A | N/A | N/A |
| Convolutional 3 | 32 | 5/2 | Same | ReLu |
| Dropout 3 | N/A | N/A | N/A | N/A |
| Batch Normalization 3 | N/A | N/A | N/A | N/A |
| Max Pooling 1 | N/A | 4/3 | Same | N/A |
| Flatten 1 | N/A | N/A | N/A | N/A |
| Fully Connected 1 | 224 | N/A | N/A | ReLu |
| Dropout 4 | N/A | N/A | N/A | N/A |
| Batch Normalization 4 | N/A | N/A | N/A | N/A |
| Output | N/A | N/A | N/A | Softmax |

**Table 8.** Table describing the architecture of the RNN model. The dropout was set to 0.35 for all instances, the batch size was 512, and the learning rate was 0.000933.

| Layer | Number of Nodes | Activation Function |
| --- | --- | --- |
| Input | 13 | ReLu |
| LSTM 1 | 96 | ReLu |
| Dropout 1 | N/A | N/A |
| Batch Normalization 1 | N/A | N/A |
| LSTM 2 | 128 | ReLu |
| Dropout 2 | N/A | N/A |
| Batch Normalization 2 | N/A | N/A |
| LSTM 3 | 256 | ReLu |
| Dropout 3 | N/A | N/A |
| Batch Normalization 3 | N/A | N/A |
| LSTM 4 | 224 | ReLu |
| Dropout 4 | N/A | N/A |
| Batch Normalization 4 | N/A | N/A |
| Fully Connected 1 | 32 | ReLu |
| Dropout 5 | N/A | N/A |
| Batch Normalization 5 | N/A | N/A |
| Output | 3 | Softmax |

### 2.8 Statistical Analysis

#### Level 1 Classification

The results were compared to identify any significant differences in the mean accuracy and mean F1 scores based on the best performing incremental learning level. Post-hoc tests, namely Wilcoxon Rank Sum test with Bonferroni corrections for multiple comparisons, were used to compare the top three performing models.

Incremental learning levels were also compared for each of the top models to determine its impact on performance. Each incremental learning level was compared to its adjacent value(s) (i.e., 0% to 10%, 10% to 20%, and 20% to 30%).

#### Offloading Data

The percentage of maximum load recorded in each position was compared for each of the three FSRs. The statistical analyses were performed twice for each sensor, for a total of six analyses, to compare all adjacent positions from 90° to 0° and from 0° to -90° (i.e., Analysis 1 was the Right Trochanter Sensor for 90° to 0°; Analysis 2 was the Right Trochanter Sensor for 0° to -90°; etc.). The data was normalized to every participant for the calculations.

#### Level 2 Classification

The results from Level 2 Classification were compared to determine the effect of bin size on model performance for right and left classification. The best classifier from each bin size was compared to its adjacent bin size(s) (i.e., 70° to 45°, 45° to 30°, and 30° to 15°).

## 3 Results

### 3.1 Participants

Descriptive statistics of the 20 participants recruited for this study are provided in Table 9.

**Table 9.** Table showing the participant demographics. *Note, participants 9 and 16 were included in this table, but they were not included in the analysis as mentioned above.

| Participant | Sex (M/F) | Age Range (years) | Height (cm) | Weight (kg) | BMI |
| --- | --- | --- | --- | --- | --- |
| 1 | F | 26-30 | 170.0 | 92.0 | 31.8 |
| 2 | M | 71-75 | 172.7 | 75.2 | 25.1 |
| 3 | M | 31-35 | 181.0 | 88.7 | 27.1 |
| 4 | M | 41-45 | 181.0 | 99.8 | 30.4 |
| 5 | F | 51-55 | 157.5 | 87.8 | 35.4 |
| 6 | F | 21-25 | 157.5 | 50.3 | 20.3 |
| 7 | M | 21-25 | 175.0 | 73.8 | 24.1 |
| 8 | M | 26-30 | 165.0 | 65.5 | 24.1 |
| 9* | F | 21-25 | 169.0 | 77.8 | 27.2 |
| 10 | M | 31-35 | 198.0 | 120.3 | 30.7 |
| 11 | F | 21-25 | 161.3 | 62.6 | 24.1 |
| 12 | F | 21-25 | 165.1 | 49.2 | 18.0 |
| 13 | F | 21-25 | 175.0 | 57.4 | 18.8 |
| 14 | F | 21-25 | 175.0 | 60.8 | 19.8 |
| 15 | F | 21-25 | 170.0 | 63.7 | 22.1 |
| 16* | F | 21-25 | 165.0 | 42.8 | 15.7 |
| 17 | M | 21-25 | 188.0 | 107.6 | 30.4 |
| 18 | M | 21-25 | 179.0 | 81.8 | 25.5 |
| 19 | M | 16-20 | 185.4 | 78.0 | 22.7 |
| 20 | M | 21-25 | 183.0 | 88.5 | 26.4 |
| Mean ± (SD) |  | 29.8 ± 13.0 | 174.4 ± 10.6 | 77.9 ± 19.2 | 25.4 ± 4.7 |

### 3.2 Level 1 Classification

Tables 10 and 11 show the overall mean accuracy and F1 scores with their respective standard deviation values across all 18 participants for the classification of supine, left, and right for each incremental learning level. Since the ILL of 30% performed best, we conducted our analyses on these models.

**Table 10.**
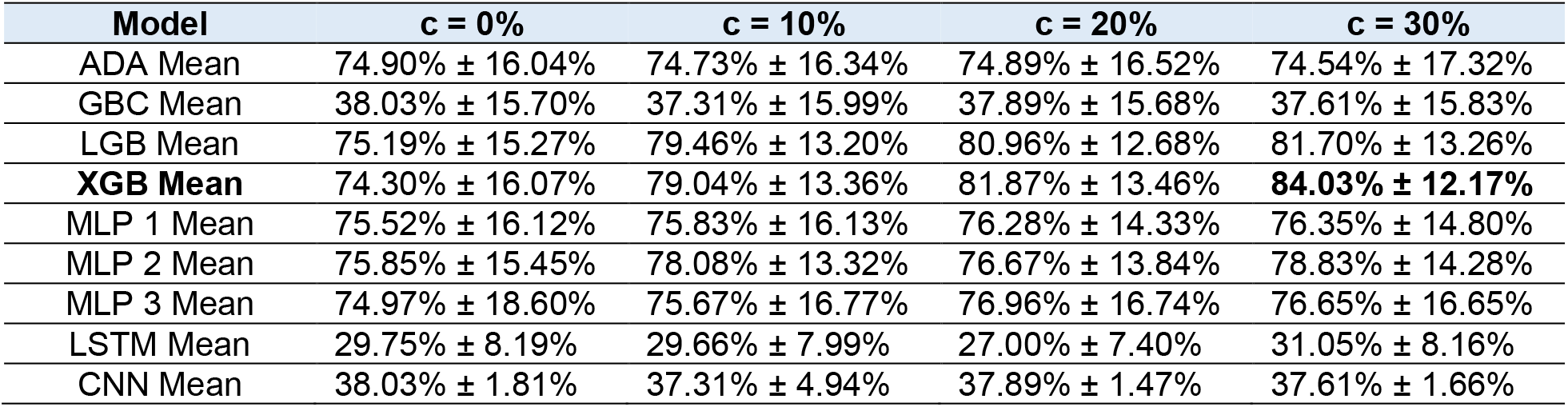
Table describing the combined mean accuracy and standard deviations of the tested models for Level 1 of the classification, which classifies positions as supine, left, or right.

| Model | c = 0% | c = 10% | c = 20% | c = 30% |
| --- | --- | --- | --- | --- |
| ADA Mean | 74.90% ± 16.04% | 74.73% ± 16.34% | 74.89% ± 16.52% | 74.54% ± 17.32% |
| GBC Mean | 38.03% ± 15.70% | 37.31% ± 15.99% | 37.89% ± 15.68% | 37.61% ± 15.83% |
| LGB Mean | 75.19% ± 15.27% | 79.46% ± 13.20% | 80.96% ± 12.68% | 81.70% ± 13.26% |
| **XGB Mean** | 74.30% ± 16.07% | 79.04% ± 13.36% | 81.87% ± 13.46% | **84.03% ± 12.17%** |
| MLP 1 Mean | 75.52% ± 16.12% | 75.83% ± 16.13% | 76.28% ± 14.33% | 76.35% ± 14.80% |
| MLP 2 Mean | 75.85% ± 15.45% | 78.08% ± 13.32% | 76.67% ± 13.84% | 78.83% ± 14.28% |
| MLP 3 Mean | 74.97% ± 18.60% | 75.67% ± 16.77% | 76.96% ± 16.74% | 76.65% ± 16.65% |
| LSTM Mean | 29.75% ± 8.19% | 29.66% ± 7.99% | 27.00% ± 7.40% | 31.05% ± 8.16% |
| CNN Mean | 38.03% ± 1.81% | 37.31% ± 4.94% | 37.89% ± 1.47% | 37.61% ± 1.66% |

**Table 11.** Table describing the combined mean F1 scores and standard deviations of the tested models for Level 1 of the classification, which classifies positions as supine, left, or right.

| Model | c = 0% | c = 10% | c = 20% | c = 30% |
| --- | --- | --- | --- | --- |
| ADA Mean | 0.7495 ± 0.1626 | 0.7478 ± 0.1662 | 0.7487 ± 0.1678 | 0.7463 ± 0.1749 |
| GBC Mean | 0.7668 ± 0.1616 | 0.7692 ± 0.1651 | 0.7680 ± 0.1625 | 0.7731 ± 0.1637 |
| LGB Mean | 0.7508 ± 0.1577 | 0.7950 ± 0.1331 | 0.8097 ± 0.1276 | 0.8176 ± 0.1331 |
| **XGB Mean** | 0.7425 ± 0.1642 | 0.7895 ± 0.1355 | 0.8181 ± 0.1356 | **0.8399 ± 0.1226** |
| MLP 1 Mean | 0.6929 ± 0.1387 | 0.6980 ± 0.1141 | 0.6762 ± 0.1697 | 0.6841 ± 0.1485 |
| MLP 2 Mean | 0.7574 ± 0.1707 | 0.7780 ± 0.1437 | 0.7665 ± 0.1467 | 0.7780 ± 0.1434 |
| MLP 3 Mean | 0.7530 ± 0.1919 | 0.7499 ± 0.2024 | 0.7691 ± 0.1756 | 0.7641 ± 0.1764 |
| LSTM Mean | 0.1423 ± 0.0673 | 0.1480 ± 0.0658 | 0.1197 ± 0.0607 | 0.1528 ± 0.0667 |
| CNN Mean | 0.2232 ± 0.0294 | 0.2216 ± 0.0302 | 0.2194 ± 0.0215 | 0.2191 ± 0.0205 |

#### Comparing Machine Learning Models

The data was confirmed to be non-parametric. A Friedman’s ANOVA reported a significant difference between the mean accuracies: χ^2^(8) = 102.15, p < 1×10^−15^ and the mean F1 scores: χ^2^(8) = 115.13, p < 1×10^−15^. Figures 5 and 6 depict the mean accuracies and F1 scores for all the models at an ILL of 30%.

**Figure 5.**
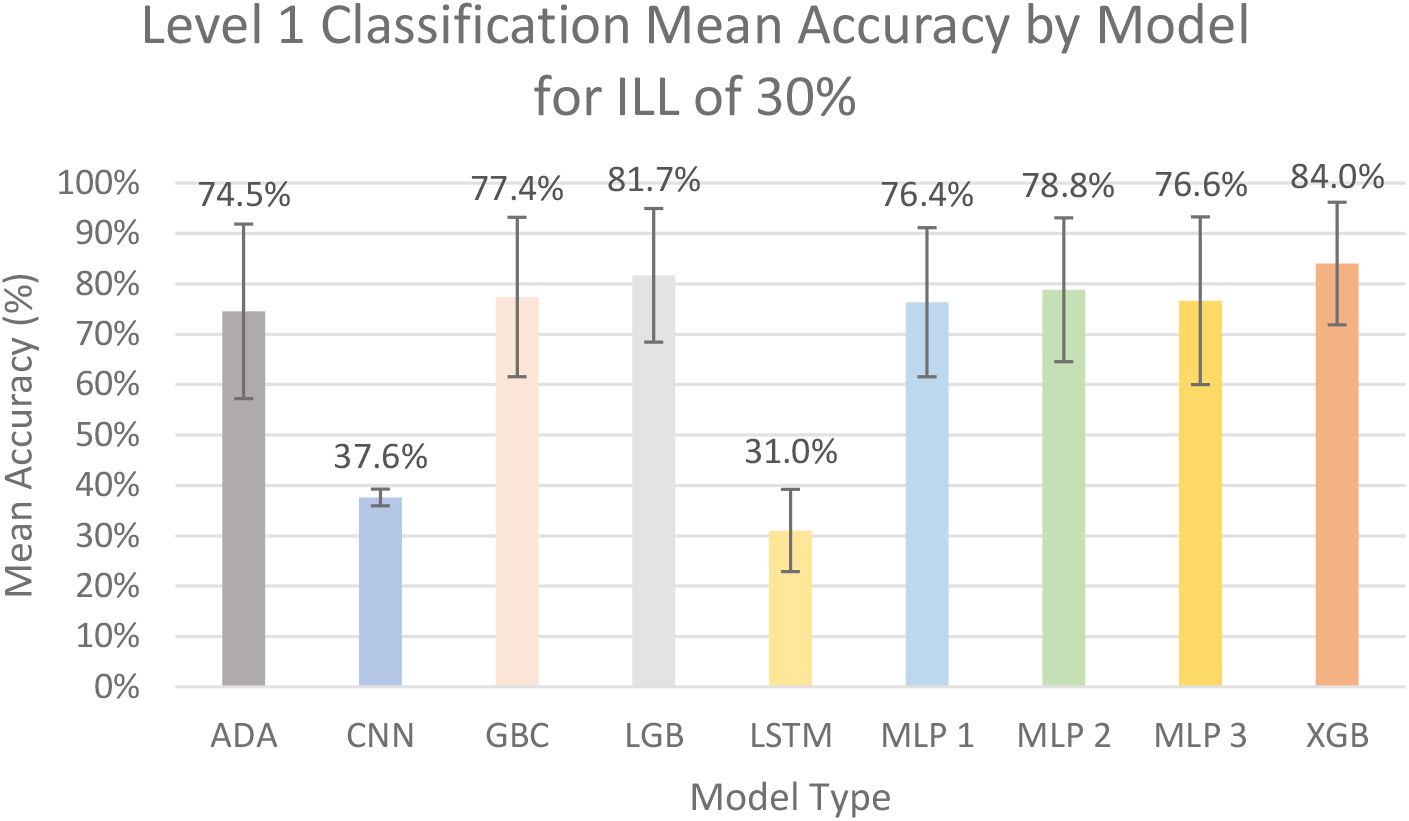
Graph of Level 1 classification mean accuracy for an ILL of 30%. The error bars represent standard deviation.

**Figure 6.**
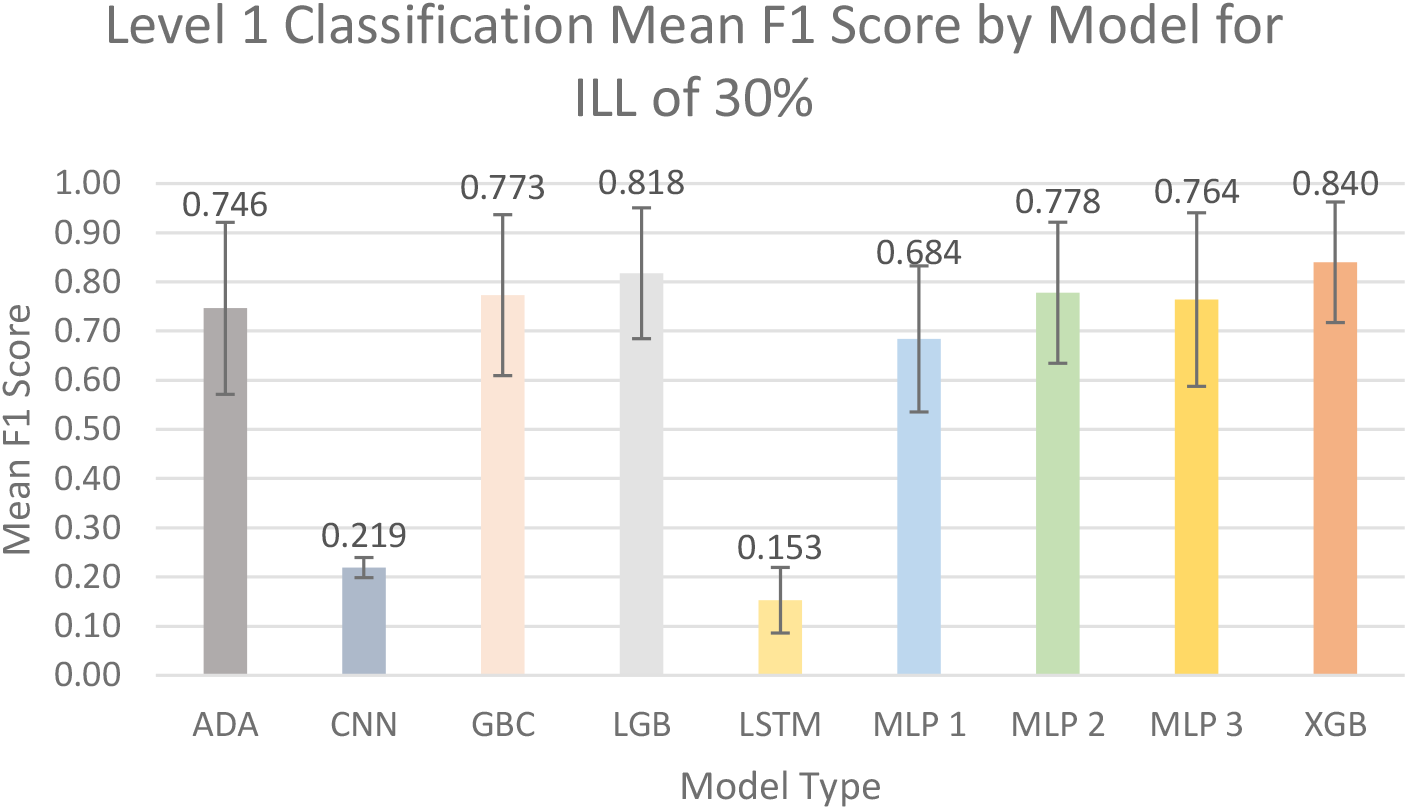
Graph of Level 1 classification mean F1 scores for an ILL of 30%. The error bars represent standard deviation.

Multiple post-hoc Wilcoxon Rank Sum tests with Bonferroni corrections were used to compare the performances of the top three models (XGB, LGB, and MLP 2). For mean accuracy, the comparison between MLP 2 vs. LGB was not significant: V = 32, p = 0.0210. The comparisons between MLP 2 vs. XGB, V = 10, p = 0.00178, and LGB vs. XGB, V = 9, p = 0.00152, were both significant. For mean F1 scores, all comparisons were found to be statistically significant, MLP 2 vs. XGB: V = 2, p = 0.000301; MLP 2 vs. LGB: V = 11, p = 0.0127; LGB vs. XGB: V = 12, p = 0.00245.

#### Comparing Incremental Learning Levels

The XGB data was confirmed to be non-parametric. A Friedman’s ANOVA reported a difference between the mean accuracies: χ^2^(3) = 37.01, p < 1×10^−7^ and the mean F1 scores: χ^2^(3) = 35.41, p < 1×10^−7^. Figure 7a and 7b show a visual comparison between the mean accuracies and F1 scores of the different incremental learning levels for the XGB model.

**Figure 7a.**
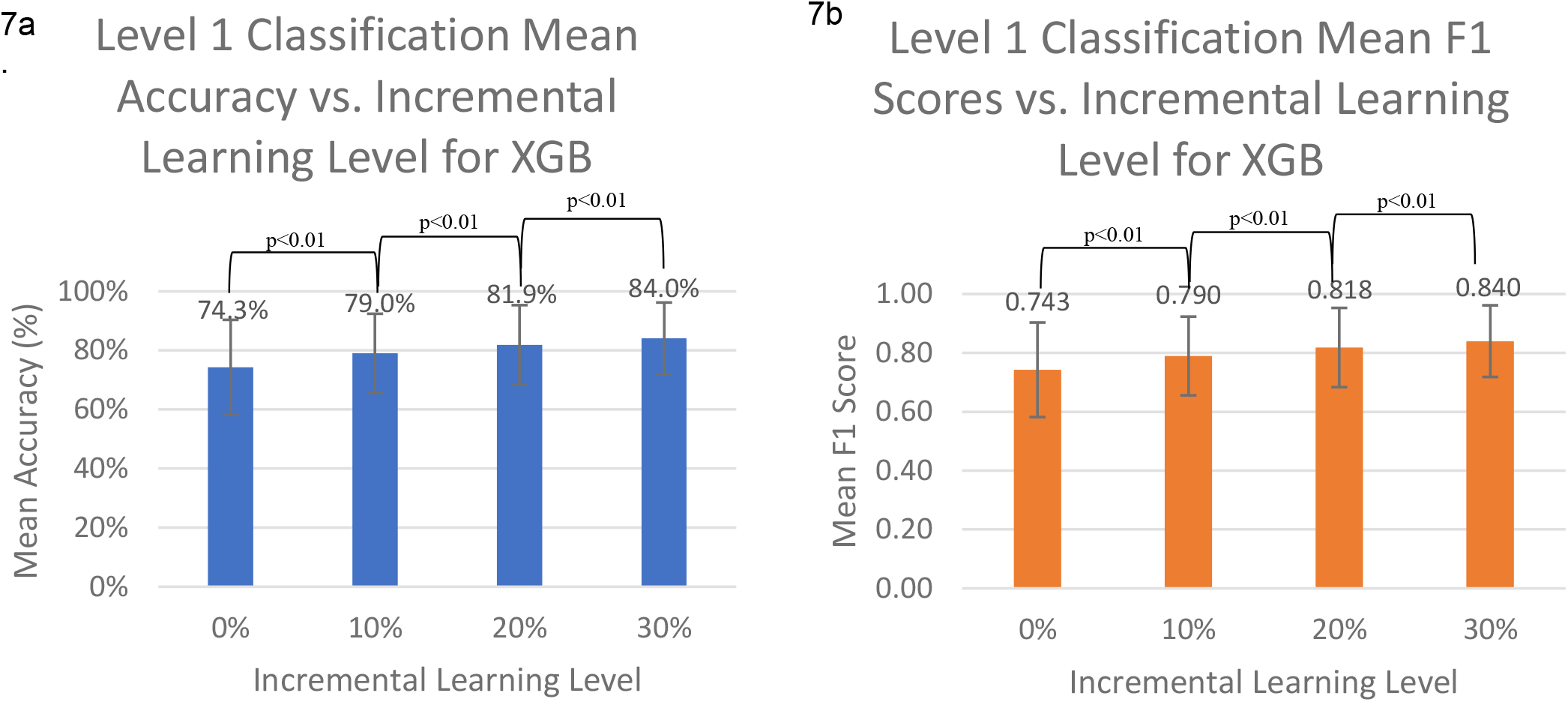
Graph of Level 1 classification mean accuracy for the top three models across the different ILLs. The error bars represent standard deviation; **7b**. Graph of Level 1 classification mean F1 scores for the top three models across the different ILLs. The error bars represent standard deviation.

Multiple post-hoc Wilcoxon Rank Sum tests with Bonferroni corrections were used to compare the XGB ILLs. All comparisons were found to be statistically significant for both mean accuracy and F1 scores. For accuracy, 0 vs. 10: V = 7, p = 0.00287; 10 vs. 20: V = 15, p = 0.00388; 20 vs 30: V = 7, p = 0.00176. For F1 scores, 0 vs. 10: V = 16, p = 0.00266; 10 vs. 20: V = 21, p = 0.00532; 20 vs. 30: V = 10, p = 0.00178.

### 3.4 FSR Data

For the left and right trochanters, only the positions from -90° to 0° and 0° to 90°, respectively, were assessed. This decision was made because a trochanter is completely offloaded when a participant is on the opposite side, meaning the force reading would be 0. The above does not hold true for the sacrum, so it was assessed for the entire range of positions from -90° to 90°.

#### Left Trochanter

The left trochanter loading data for positions -90° to 0° was analyzed and confirmed to be non-parametric. A Friedman’s ANOVA was significant, χ2(5) = 71, p < 1×10^−13^.

Multiple post-hoc Wilcoxon Rank Sum tests with Bonferroni corrections were used to compare adjacent primary positions to determine if there was a difference in percentage of maximum load. In total, five comparisons were made, changing the p-value needed to reach significance to p < 0.01. Two of the comparisons were statistically significant and two almost reached statistical significance, -90° to -60°: V = 106, p = 0.051; -60° to -45°: V = 132, p < 0.001; -45° to -30°: V = 22; p = 0.016; -30° to -15°: V = 136, p < 1×10^−4^; -15° to 0°: V = 116, p = 0.011.

Figure 8 shows the percentage of max load felt at the left trochanter as participants rotated through different positions and the significance between positions.

**Figure 8.**
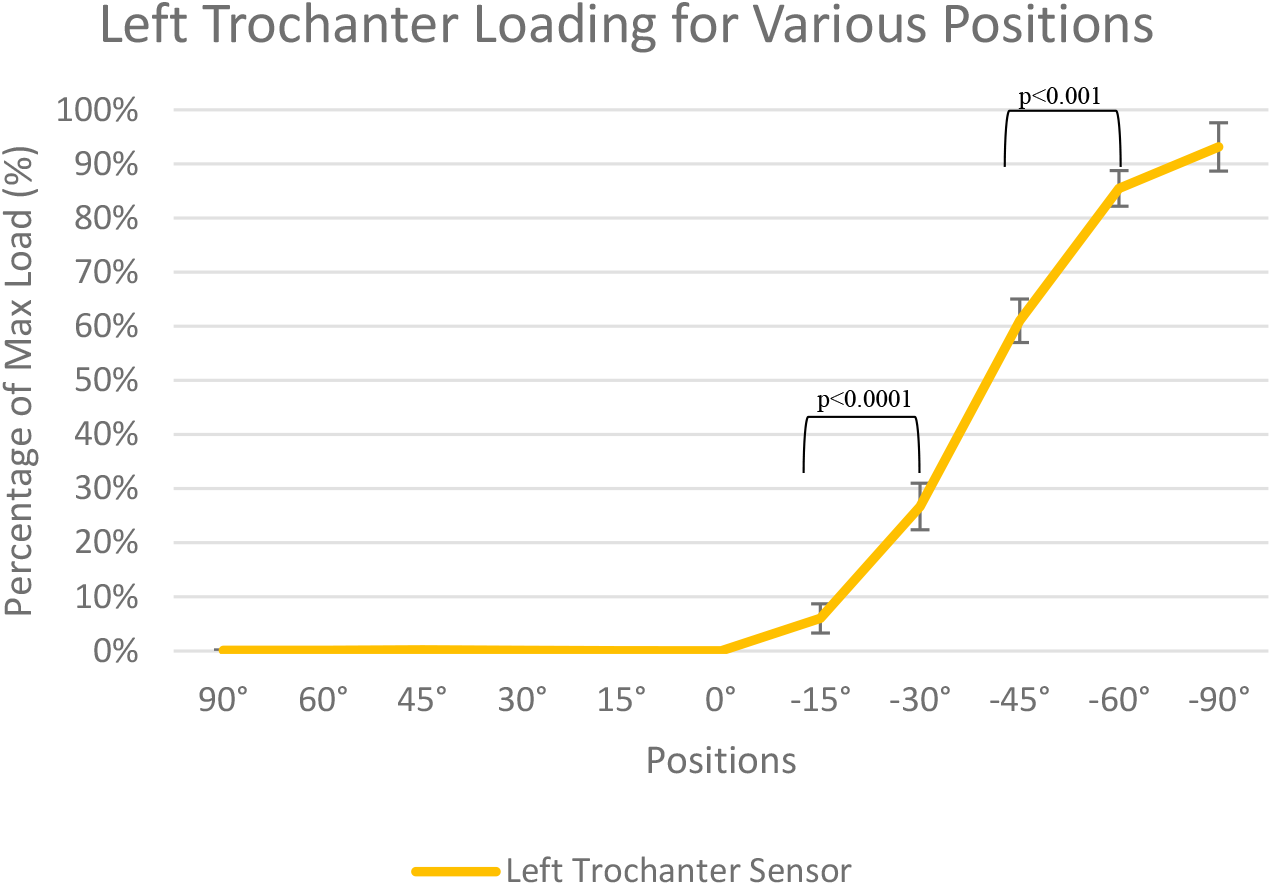
Graph of the percentage of maximum load for the left trochanter sensor for all the primary positions. The error bars represent the standard error.

#### Right Trochanter

The right trochanter loading data for positions 90° to 0° was analyzed and confirmed to be non-parametric. A Friedman’s ANOVA was significant, χ2(5) = 78.48, p < 1×10^−14^.

Multiple post-hoc Wilcoxon Rank Sum tests with Bonferroni corrections were used to compare adjacent primary positions to determine if there was a difference in percentage of maximum load. In total, five comparisons were made, changing the p-value needed to reach significance to p < 0.01. All comparisons except from 90° to 60° were statistically significant, 90° to 60°: V = 117, p = 0.057; 60° to 45°: V = 152, p < 1×10^−4^; 45° to 30°: V = 146; p < 0.001; 30° to 15°: V = 153, p < 1×10^−4^; 15° to 0°: V = 140, p < 0.01.

Figure 9 shows the percentage of max load felt at the right trochanter as participants rotated through different positions and the significance between positions.

**Figure 9.**
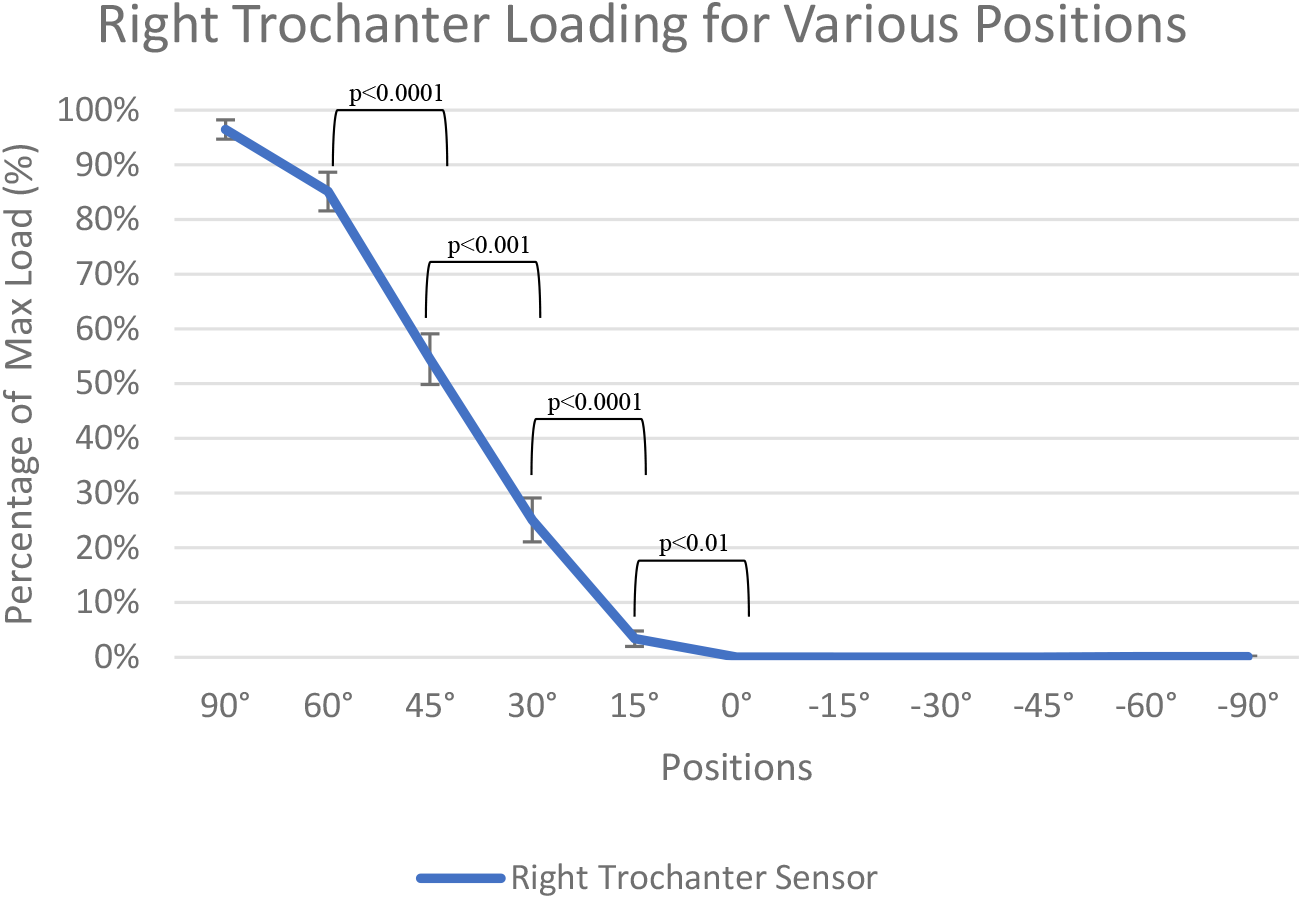
Graph of the percentage of maximum load for the right trochanter sensor for all the primary positions. The error bars represent the standard error.

#### Sacrum

The sacral loading data for positions 90° to 0° was analyzed and confirmed to be non-parametric. A Friedman’s ANOVA was significant, χ2(5) = 75.218, p < 1×10^−14^.

Multiple post-hoc Wilcoxon Rank Sum tests with Bonferroni corrections were used to compare adjacent primary positions to determine if there was a difference in percentage of maximum load. In total, five comparisons were made, changing the p-value needed to reach significance to p < 0.01. Four of the comparisons were statistically significant, 90° to 60°: V = 7, p < 0.001; 60° to 45°: V = 7, p < 0.001; 45° to 30°: V = 55; p = 0.33; 30° to 15°: V = 4, p < 0.001; 15° to 0°: V = 9, p < 0.001.

The sacral loading data for positions -90° to 0° was analyzed and confirmed to be non-parametric. A Friedman’s ANOVA was significant, χ2(5) = 67.679, p < 1×10^−12^.

Multiple post-hoc Wilcoxon Rank Sum tests with Bonferroni corrections were used to compare adjacent primary positions to determine if there was a difference in percentage of maximum load. In total, five comparisons were made, changing the p-value needed to reach significance to p < 0.01. Three of the comparisons were statistically significant and one almost reached statistical significance, -90° to -60°: V = 10, p < 0.01; -60° to - 45°: V = 60, p = 0.71; -45° to -30°: V = 22; p = 0.016; -30° to -15°: V = 1, p < 1×10^−4^; - 15° to 0°: V = 13, p = 0.0027.

Figure 10 shows the percentage of maximum load felt at the sacrum as participants rotated through different positions and the significance between positions.

**Figure 10.**
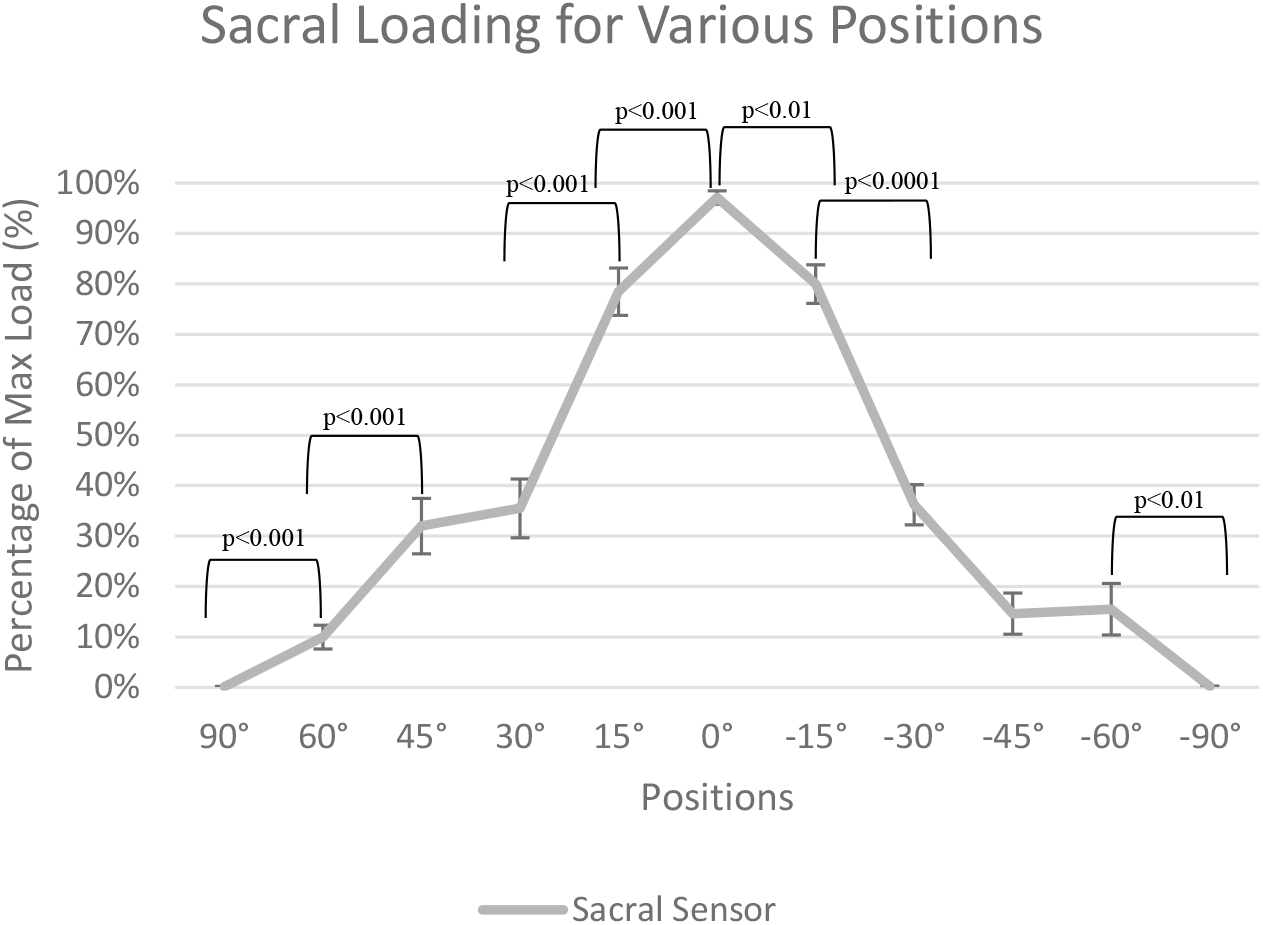
Graph of the percentage of maximum load for the sacral sensor for all the primary positions. The error bars represent the standard error.

### 3.5 Level 2 Classification

Tables 12 and 13 show the overall mean accuracy and F1 scores with their respective standard deviation values across all 18 participants from Level 2 left and right classification using an ILL of 30%. Only the top two models are shown.

**Table 12.**
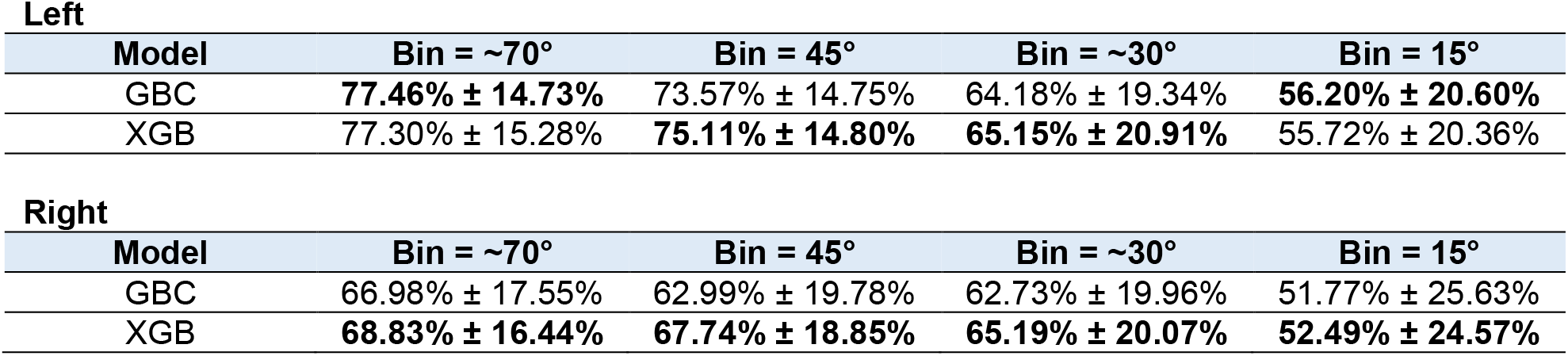
Table showing the mean accuracy and standard deviation values for the Level 2 Right and Left Classifications of different bin sizes for the top two models.

**Table 13.** Table showing the mean F1 scores and standard deviation values for the Level 2 Right and Left Classifications of different bin sizes for the top three models.

| Left |  |  |  |  |
| --- | --- | --- | --- | --- |
| Model | Bin = ~70° | Bin = 45° | Bin = ~30° | Bin = 15° |
| GBC | 0.7080 ± 0.1963 | 0.6907 ± 0.1855 | 0.6031 ± 0.2179 | 0.5425 ± 0.2022 |
| XGB | 0.7159 ± 0.2011 | 0.7100 ± 0.1864 | 0.6213 ± 0.2312 | 0.5400 ± 0.2099 |

| Right |  |  |  |  |
| --- | --- | --- | --- | --- |
| Model | Bin = ~70° | Bin = 45° | Bin = ~30° | Bin = 15° |
| GBC | 0.5914 ± 0.2299 | 0.5712 ± 0.2285 | 0.5780 ± 0.2224 | 0.4886 ± 0.2566 |
| XGB | 0.6097 ± 0.2239 | 0.6181 ± 0.2305 | 0.6039 ± 0.2270 | 0.4996 ± 0.2573 |

#### Comparing Bin Sizes

The data for Level 2 left classification was confirmed to be parametric for both mean accuracy and F1 scores. Two ANOVAs reported a significant difference between the different bin sizes for mean accuracies, F(68,3) = 5.004, p = 0.0034, and no significant difference for the mean F1 scores, F(68,3) = 2.714, p = 0.0516. Figures 11a and 11b show the best mean accuracies and F1 scores from each Level 2 right classification bin.

**Figure 11a.**
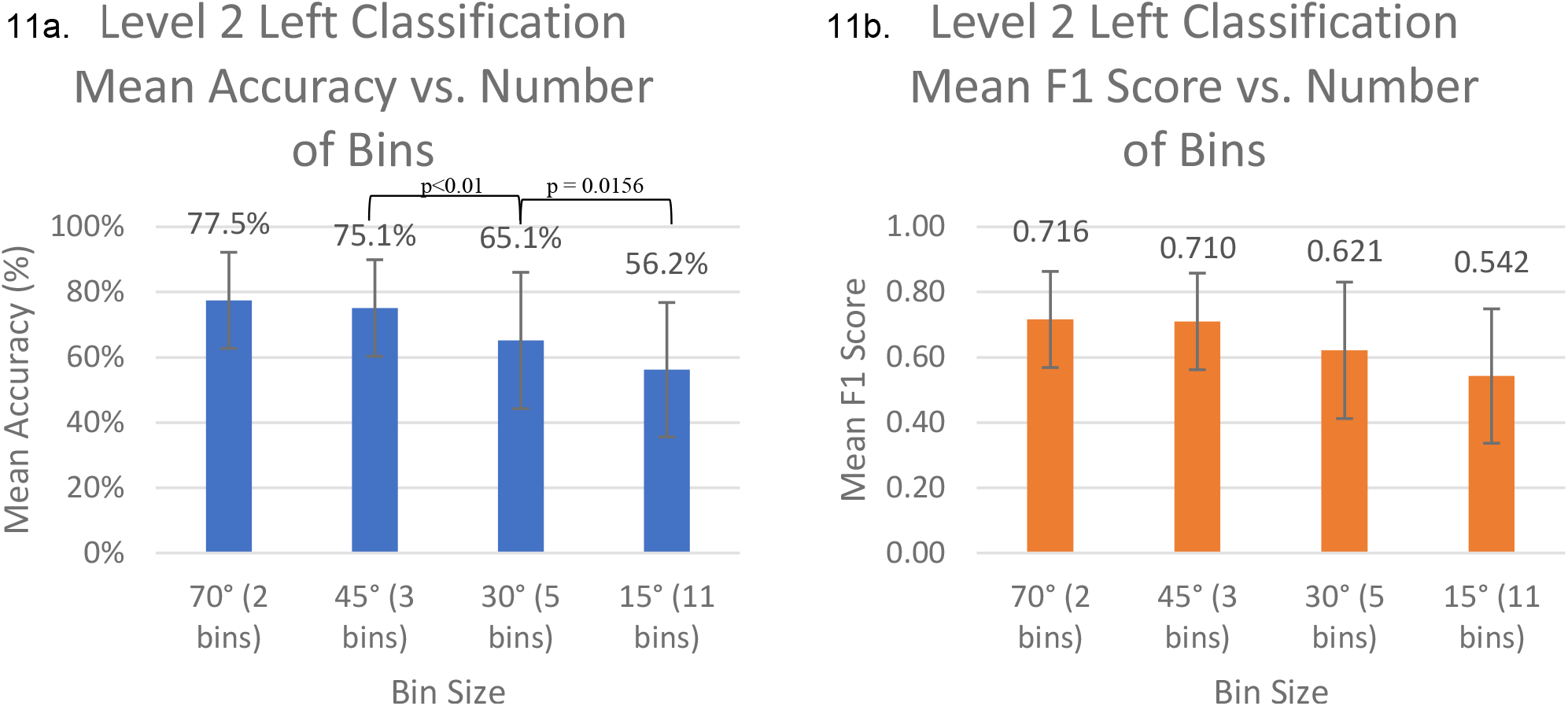
Graph of mean accuracy for the top models for Level 2 left classification for the four different bin sizes; **11b**. Graph of mean F1 score for the top models for Level 2 left classification for the four different bin sizes.

Multiple post-hoc t-tests with Bonferroni corrections were used to compare the adjacent bin sizes. In total, three comparisons were made, changing the p-value needed to reach significance to p < 0.017. For mean accuracy, the 45° vs 30° and 30° vs 15° bin comparisons were found to be statistically significant: 45° vs 30°: t(17) = 2.954, p = 0.00889; 30° vs 15°: t(17) = 2.685, p = 0.0156, respectively.

The data for Level 2 right classification was confirmed to be parametric for both mean accuracies and F1 scores. Two ANOVAs reported no significant difference between the different bin sizes for mean accuracies, F(68,3) = 2.369, p = 0.0782, and F1 scores, F(68,3) = 0.959, p = 0.417. Figures 12a and 12b show the best mean accuracies and F1 scores from each Level 2 right classification bin. No post-hoc tests were performed.

**Figure 12a.**
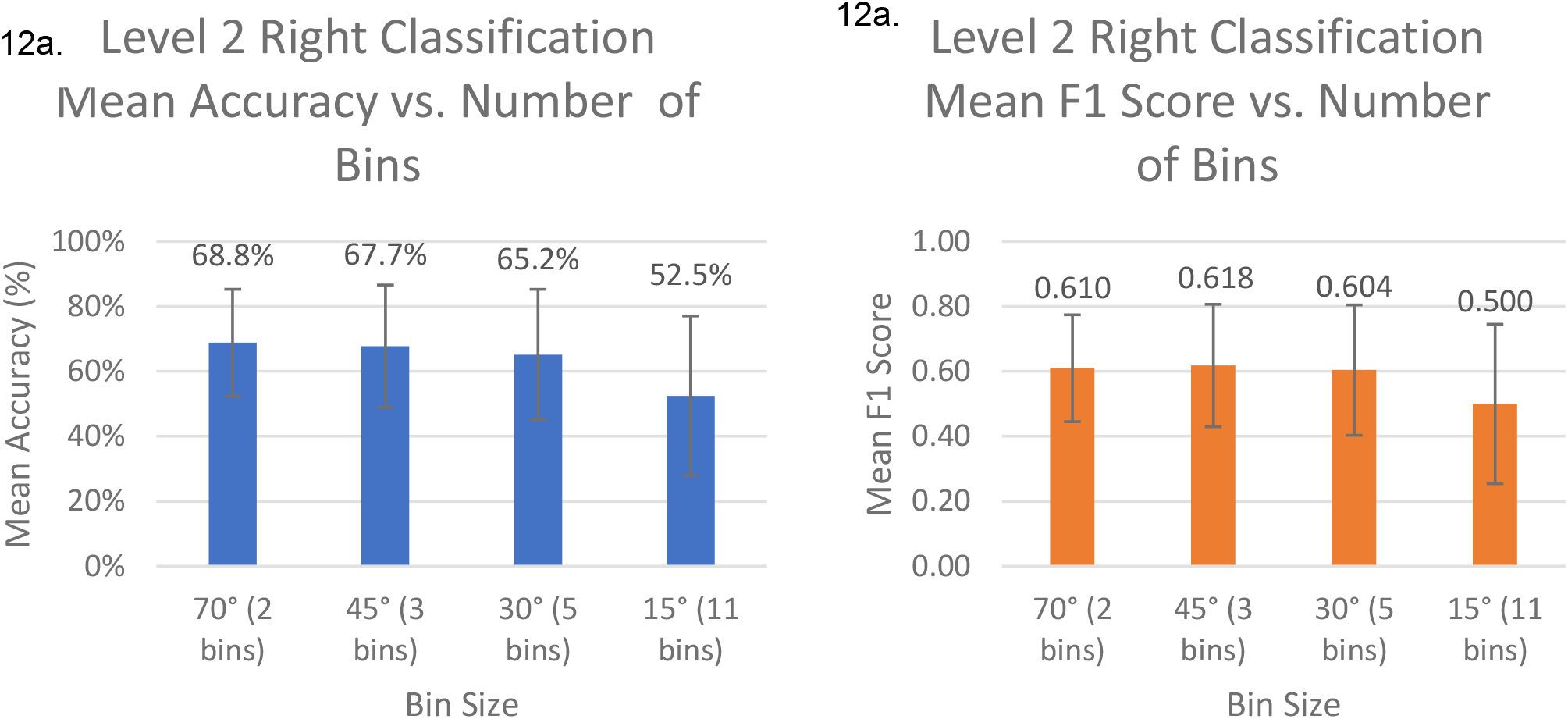
Graph of mean accuracy for the top models for Level 2 left classification for the four different bin sizes; **12b**. Graph of mean F1 score for the top models for Level 2 left classification for the four different bin sizes.

## 4 Discussion

### 4.1 Level 1 Classification

The best performing model was the XGBoost model with a mean accuracy of 84.03% ± 12.17%, and mean F1 score of 0.8399 ± 0.1226. This accuracy is an improvement over the ∼68% that Wong et al.’s previous model was able to achieve on this data. It is likely that this accuracy represents an underestimate of the actual accuracy of our new system as the training set includes data without the IMU ground truth data.

The deep learning methods generally performed worse than the machine learning models, except for the MLP 2 model which performed at a comparable level to the LGB. However, this model had an overly complicated architecture for the amount of data it was processing, so it may have overfit to the data. In general, the most likely reason for the poor performance of the CNN and RNN was the lack of data as deep learning traditionally relies on very large data sets.

#### Comparing Machine Learning Models

The omnibus test found an overall significant difference between the performance of all models and post-hoc comparisons of the top three models found statistically significant differences. In terms of mean accuracy and F1 score, the XGB model performed significantly better than both the LGB and MLP 2 models. For mean accuracy, the LGB and MLP 2 were found to have performed comparably, whereas for mean F1 score, the LGB model outperformed the MLP 2 model. Therefore, the XGB model was statistically significantly better than the other models and should thus be included in future work when testing position prediction.

#### Comparing Incremental Learning Levels

A Friedman’s ANOVA showed a significant difference between the performance of the XGB model at different ILLs. Post-hoc tests further confirmed that there was a significant improvement in performance between adjacent ILLs for both mean accuracy and F1 score as the ILL increased. This finding is important as it suggests that collecting data for incremental learning has the potential to better personalize the model to participants, thus improving their care. It is important to note that the test set for each participant in the study was limited ranging from 83 to 215 observations (mean 151.1 observations), meaning that a maximum of 64 observations at 30% incremental learning were added to a data pool of ∼20,000 observations for incremental learning. Considering that incremental learning with so few observations was able to improve the overall mean accuracy and F1 score by almost 10% and 0.1, respectively, for the XGB model, it would be important to further investigate the impacts of incremental learning with a larger data set. Additionally, it would be important to investigate whether the statistical significance of incremental learning translates to clinical significance and an improved prevention of PIs.

### 4.2 FSR Data

#### Left Trochanter

The left trochanter was loaded for most angles between 0° and -90° (as shown in Figure 8). A Friedman’s ANOVA indicated that there was a significant difference between the mean percentage of maximum force experienced on the left trochanter between different left-side lying positions. Post-hoc comparisons compared adjacent left positions and identified that the percentage of maximum force on the left trochanter was significantly different for -60° to -45° and -30° to -15°. Changes from -45° to -30° and -15° to 0° almost met significance. This finding indicates that rotating a participant from a more extreme position to a less extreme position between the positions -60° to -45° and -30° to -15° will result in significant offloading of the left trochanter compared to the previous position.

#### Right Trochanter

The right trochanter was loaded for most angles between 0° to 90° (as shown in Figure 9). A Friedman’s ANOVA indicated that there was a significant difference between the mean percentage of maximum force experienced on the right trochanter between different right-side lying positions. Post-hoc comparisons compared adjacent right positions and identified that the percentage of maximum force on the right trochanter was significantly different for all adjacent positions except for 90° to 60°. This finding indicates that rotating a participant from a more extreme position to a less extreme position between the positions 60° to 45°, 45° to 30°, 30° to 15°, and 15° to 0° will result in significant offloading of the right trochanter compared to the previous position.

#### Sacrum

The sacrum was only completely offloaded at -90° and 90° (as shown in Figure 10). A Friedman’s ANOVA reported a significant difference between the mean percentage of maximum force experienced on the sacrum between different right-side lying positions. Post-hoc comparisons compared adjacent right positions and identified that the percentage of maximum force on the sacrum was significantly different for all adjacent positions except for 45° to 30°. This finding indicated that rotating a participant from a less extreme position to a more extreme position between the positions 0° to 15°, 15° to 30°, 45° to 60°, and 60° to 90° resulted in significant offloading of the sacrum compared to the previous position.

A Friedman’s ANOVA reported a significant difference between the mean percentage of maximum force experienced on the sacrum between different left-side lying positions. Post-hoc comparisons were used to compare adjacent left positions and they identified that the percentage of maximum force on the sacrum was significantly different for -90° to -60°, -30° to -15°, and -15° to 0°. Changes from -45° to -30° almost met significance. This finding indicates that rotating a participant from a less extreme position to a more extreme position between the positions 0° to -15°, -15° to -30°, and -60° to -90° will result in significant offloading of the sacrum compared to the previous position.

#### Optimal Offloading

The trochanter opposite to the side a patient was turned on will be completely offloaded, making the trochanters easier to offload than the sacrum. It appeared that the sacrum was not fully offloaded in any position that required the use of a support pillow as the patient’s sacrum was likely pressed up against it. If complete offloading is required for adequate tissue healing, it may be necessary for patients to assume a side-lying position that can be maintained without the use of assistive device to maintain the position. If assistive devices are needed, it may be important to ensure they have a cut out around the sacral area to ensure it is being properly offloaded.

#### Optimal Bin Size

The results indicated that the smallest bin size needed to detect meaningful changes in offloading is 15°. However, if patients require complete offloading to heal, then classifying positions as supine, left, or right will suffice. As such, it may be more important to focus on accurately detecting large positional changes like those in Level 1 classification to ensure offloading is occurring on a scheduled basis. More precise detection may be useful in recognizing smaller self-repositioning efforts and determining their impact on high-risk areas.

### 4.3 Level 2 Classification

Level 2 classification separates the *right, left*, and *supine* classifications from Level 1 classification into smaller bins. Tables 12 and 13 summarize the results from the top two models for Level 2 classification of left and right bins based on bin size. The best performing models vary depending on bin size, but the XGB model was best in six out of eight cases for mean accuracy and seven out of eight for mean F1 score. The table also shows a trend of left-side positions being predicted correctly more often than right-side positions. The reason for this finding is currently unclear.

#### Comparing Bin Sizes

Tables 12 and 13 and Figures 11 and 12 show the effect of bin size on prediction accuracy. The results show that the accuracy of predictions decreases as the precision, or number of bins, increases.

ANOVAs indicated that the only significant difference in bin sizes was in the Level 2 left mean accuracy comparison. When further analyzed, post-hoc t-tests indicated that the 45° vs 30° and 30° vs 15° bin comparisons were significant.

These results are important because they indicate that there is likely a trade-off between accuracy and precision when making predictions. It will be important to optimize the bin size for this system to ensure it is recording and classifying movements of interest. In the future, bin size should be optimized based on information gathered from offloading data and clinical expertise to decide what is the smallest positional change that needs to be captured.

### 4.4 Study Limitations

This study included a number of technical and clinical limitations as described below:

The technical limitations of this study included:

1. Load cell data was filtered with customized parameters for each participant, which may have led to an increase in classification accuracy compared to using generic parameters.
2. Most of the training data did not contain IM ground truth, so it could have negatively impacted the accuracy.
3. The data set was too small to run one-shot learning to compare its accuracy with the current hierarchical approach.
4. The neural network architectures selected by hyperparameter tuning were very complicated and may have overfit the data.
5. Level 2 classification had an imbalance of positions that were >90° and <-90°, which could have negatively impacted the prediction accuracy for more extreme positions.

The clinical limitations of this study included:

1. The patient population was predominantly young, healthy individuals, which likely did not reflect the performance with a population of older adults with/at-risk of PI.
2. Certain primary positions participants were asked to adopt were unnatural, which may have impacted the participants’ abilities to relax/breathe normally.
3. Individuals were supported using pillows that were occasionally placed against the sacrum in side-lying positions, which could have resulted in overestimates in sacral load. Train clinicians would have likely avoided placing the support pillows against the sacrum.
4. The way in which the IMUs were attached could have been more reliable to ensure they did not move while patients were changing positions.
5. The FSRs were placed by participants under guidance of the author, which means they may not have been placed in the correct anatomical position every time.
6. The FSRs occasionally fell off the participants during the study and needed to be re-attached mid study.

### 4.5 Future Work

Future work will include:

1. Collecting overnight data from at-risk patients in their own homes.
2. Revising pillow placement during offloading and trying out different repositioning aids to see the impacts on sacral loading.
3. Validate a safe way to use IMUs for overnight data collection of at-risk patients.
4. Treating the detection of patient position as a regression task instead of a classification task to evaluate performance.

## 5 Conclusion

The main findings of this study were:

1. An IMU mounted to the pelvis improved position detection accuracy for supine, left, or right from ∼70% in our previous work to 84.2% ± 11.8% for the best performing model.
2. The right and left trochanters were completely offloaded for TPAs of 0° to 90° and 0° to -90°, respectively. The sacrum was only completely offloaded for TPAs of >=90° and <=-90°, highlighting a potential limitation of the existing clinical guidelines suggesting individuals be rotated between TPAs of -40° and 40°.
3. Prediction accuracy decreased as the precision increased.

## Data Availability

All data produced in the present study are available upon reasonable request to the authors.

